# The impact of COVID-19 on acute Trauma and Orthopaedic referrals and surgery in the UK: the ‘golden peak weeks’ of the first national multi-centre observational study: The COVid-Emergency Related Trauma and orthopaedics (COVERT) Collaborative

**DOI:** 10.1101/2020.08.22.20179770

**Authors:** Kapil Sugand, COVERT Collaborative

## Abstract

**Objectives:** This is the first national study observing the impact of the COVID-19 pandemic on orthopaedic trauma with respect to referrals, operative caseload and mortality during the first six weeks (namely the “golden peak weeks”) following the introduction of the national social distancing and lockdown measures from mid-March 2020.

**Design:** A longitudinal, national, multi-centre, retrospective, observational, cohort study was conducted for the first six weeks from March 17, 2020 from start of the national social distancing and then lockdown compared to the same period in 2019 as a comparative baseline.

**Setting:** Hospitals from seven major urban cities were recruited around the UK, including London, representing a comprehensive national picture of the impact of COVID-19 pandemic and its lockdown at its peak.

**Participants:** A total of 4840 clinical encounters were initially recorded. Exclusion criterion consisted of spinal pathology only. Post-exclusion, 4668 clinical encounters were recorded and analysed within the two timeframes.

**Main outcome measures:** Primary outcomes included the number of acute trauma referrals and those undergoing operative intervention, patient demographics, mortality rates, and the proportion of patients contracting COVID-19. Secondary outcomes consisted of the mechanism of injury, type of operative intervention and proportion of aerosolising-generating anaesthesia utilised. Demographics for each patient was recorded along with underlying medical co-morbidities. Sub-group analysis compared mortalities between both cohorts. Statistical analyses included mean (±SD), risk and odds ratios, as well as Fisher’s exact test to calculate the statistical significance (p≤0.05).

**Results:** During the COVID-19 period there was a 34% reduction in acute orthopaedic trauma referrals compared to 2019 (1792 down to 1183 referrals), and 29.5% less surgical interventions (993 down to 700 operations). The mortality rate significantly (both statistically and clinically) more than doubled for both risk and odds ratios during the COVID period in all referrals (1.3% vs 3.8%, p = 0.0005) and in those undergoing operative intervention (2.2% vs 4.9%, p = 0.004). Moreover, mortality due to COVID-related complications (versus non-COVID causes) had greater odds by a factor of at least 20 times. The odds ratios of road traffic accidents, sporting injuries, infection, and lower limb injuries were significantly less (by a third to a half) during the COVID period; albeit, the odds of sustaining neck of femur fractures and having falls < 1.5m increased by more than 50%.

For the operative cohorts, there was a greater odds of aerosolising-generating anaesthesia (including those with superimposed regional blocks) by three-quarters as well as doubling of the odds of a Consultant acting as the primary surgeon. Nevertheless, the odds of open reduction and internal fixation reduced by a quarter whereas removal of metalwork or foreign bodies reduced by three-quarters. Six-week Kaplan-Meier survival probability analysis confirmed those patients with neck of femur fracture and pre-existing cardiovascular and cerebrovascular disease were most at risk of mortality during the COVID-19 era.

**Conclusion:** Although there was a reduction of acute trauma referrals and those undergoing operative intervention, the mortality rate still more than doubled in odds during the peak of the pandemic compared to the same time interval one year ago. Elderly patients with neck of femur fractures and existing cardiovascular and cerebrovascular comorbidities were at the highest risk stratification for mortality. This was the first national study to assess impact of COVID-19 pandemic on acute Orthopaedic trauma and it will aid clinicians in counselling trauma patients of the increased risk of mortality during the era of COVID-19 as well as acting as a risk-prediction tool influencing policymaking as the pandemic continues with potential subsequent waves. Further studies after the lifting of the lockdown are also required to observe for return of standard practice.

## Introduction

### The Global Impact of COVID-19

The COVID-19 pandemic will be remembered as one of the most unprecedented global health crises in modern history. With almost 700,000 deaths and 17 million recorded cases globally, the pandemic has had a permanent impact on healthcare at the time of article submission.^1^ The viral outbreak was first reported in December 2019 with the first patient hospitalised in the city of Wuhan, China.^2^ By mid-March the outbreak affected over 190 countries with over 450,000 cases and over 20,000 deaths, thus being declared a pandemic and a global public health emergency by the World Health Organization.^3^ On January 24^th^ 2020 Europe reported its first case followed by a case in the United Kingdom (UK) 5 days later.^4^

### The British Response to the pandemic

The English government responded by implementing social distancing measures on the 17^th^ March 2020 in an attempt to reduce the rate of transmission and therefore the demands on the National Health Service (NHS).^5^ This was followed a week later by more stringent measures, commonly referred to as a societal ‘lockdown’.^6^ As of the 23^rd^ March 2020, all members of the public were required to stay at home. The NHS has also been deeply affected by the strain imposed by the virus as the healthcare infrastructure has had to evolve to cope with the overwhelming and unexpected pressures on staff, resources and finances. There has been a complete renovation of Emergency Medicine and Orthopaedic services to manage musculoskeletal disease and trauma. In response to the NHS emergency declaration,^7^ the Royal Colleges of Surgeons^8^ and the British Orthopaedic Association^9^ both issued statements and guidelines for delivering emergency Trauma and Orthopaedic care during the COVID-19 outbreak. The lockdown to limit the spread of the virus has had an unforeseen effect in significantly reducing the acute trauma workload described in several single centre studies with reductions reported between 33-50%.^10,11^ There has however not been a national reflection of the impact of the COVID-19 pandemic on the orthopaedic workload and its potential impact on the mortality.

## Aim

To observe the impact of COVID-19 on Trauma and Orthopaedic acute referrals, operative casemix and mortality rates during the ‘golden peak weeks’ of the pandemic compared to the same time interval in 2019.

## Outcomes

Primary outcomes included the number of acute trauma referrals and those undergoing operative intervention, patient demographics, mortality rates, and the proportion of patients contracting COVID-19. Secondary outcomes consisted of the mechanism of injury, type of operative intervention and proportion of aerosolising-generating anaesthesia utilised.

## Alternative hypothesis

The alternative hypothesis was that when comparing both years, there would be a difference in the prevalence of acute orthopaedic referrals, orthopaedic trauma casemix and aerosol-generating anaesthetic procedures due to social distancing/lockdown. Mortality rates and survival probabilities were also hypothesised to differ due to COVID-19 outbreak.

## Methods

### Study design

This is the first and the largest national multi-centre longitudinal observational study observing patients who were acutely referred to the Trauma and Orthopaedic departments as well as those operated on within the same six-week interval comparing 2019 to 2020.

### Patient sampling

All acute referrals, operative notes, inpatient medical records and discharge
summaries were accessed using electronic medical system at each contributing hospital trust.

### Patient and Public Involvement

Although there was no active involvement of patients or the public for the study, the public body was kept in mind when conducting this observational study. Having been thrust with little preparation and notice into an unprecedented global pandemic, the collaborative authors have all been recruited to work at grass root level to combat the disease burden. We have strived as a collective to research on the effect of the COVID-19 virus on our versatile range of patients presenting with musculoskeletal injuries or requiring urgent and emergent Orthopaedic surgery. This study was set up for the patients who have been ravaged by the devastating respiratory illness so that acute and emergency services may learn to evolve alongside the virus. Information was gathered from patient records as part of auditing departmental activity. Rest assured, all data were treated confidentially and anonymity was strongly upheld throughout the study. This study was intended to influence policymaking as the pandemic continues across the UK with subsequent waves. If the pandemic continues to threaten the lives of those vulnerable, Orthopaedic surgeons and patients may be swayed to avoid surgery and anaesthetic intubation altogether. Therefore, those within a high-risk stratification may be managed conservatively and non-operatively as long as the risks and the benefits of surgery have been thoroughly discussed and a mutual agreement between patient and surgeon has been reached. There are certain procedures which do require surgery sooner rather than later, but if the parameters of the deformity or injury are within acceptable limits, then surgery may be delayed or avoided as long as there is no functional deficit. Decision-making in surgery ought to be conducted by both parties alongside multi-disciplinary input to provide optimal and holistic patient-centred care. Our tireless efforts are dedicated to all those who have been affected by this plague.

### Study period

The six-week study period was from the start of social distancing on Tuesday 17^th^ March 2020 to Tuesday 31^st^ April 2020 which encompassed the national ‘lockdown’ measures instigated on the 23^rd^ March 2020. This period was considered the ‘golden peak’ of the epidemic in the UK. This was compared to the same six-week interval from Tuesday 19^th^ March to Tuesday 30^th^ April 2019 (i.e. prior to any COVID-19 related measures) to compare the impact of the pandemic one year apart.

### Inclusion criteria

All acute orthopaedic trauma referrals presenting to the Emergency Department during the intervals one year apart were included. All orthopaedic trauma cases that required an operation, including those from acute orthopaedic trauma referrals, within the intervals one year apart. Those patients listed for an operation prior to time period of data collection were included in the final analysis. We adhered to STROBE guidelines for observational studies.

### Exclusion criteria

Any cases being referred internally from other specialties for Trauma and Orthopaedic advice and input, as well as referrals from any external centre asking for tertiary advice were excluded from further analysis. Any patients with post-operative complications arising from the period prior to the data collection were excluded. For operative trauma cases, those undergoing spinal procedures were excluded as these are jointly treated by Neurosurgery in most hospitals. All non-urgent semi-elective procedures were excluded from analysis as well, as they would inaccurately assess the impact of any social distancing measures on the trauma workload. Routine elective orthopaedic cases were excluded.

### Data points

Demographics including age, sex and ASA grades were recorded for all patients. Injury characteristics were recorded, including the anatomical location and if the injury was open or closed. The mechanism of injury was categorised and whether the patient was referred as a trauma call. The nature of the operative procedures and the anaesthetic techniques were recorded. Patients undergoing multiple procedures were recorded for every episode where they were taken to theatre. Six-week mortality rate was recorded as well as the COVID-19 status of any symptomatic patients or suspected cases. Data points were divided into acute referrals and operative casemix as seen in table 1.

- *Anaesthetic techniques:* This was divided into anaesthetic aerosolising-generating procedures (AGP) which consisted of any intubation (including laryngeal mask airway and endotracheal intubation) for a general anaesthetic. All other anaesthetic techniques including regional and local anaesthetics were deemed as non-AGPs.
- *COVID status*: groups of patients were divided into either not swabbed, swabbed due to presence of documented symptoms, negative swabs and positive results.

**Table 1:** data points for acute referrals and operative casemix

|  |  |  |  |
| --- | --- | --- | --- |
| Age (years) | Gender (Male / female by birth) | ASA (1-5) | Date of injury / presentation |
| Injury | Mechanism of injury | Open Vs Closed fracture | Trauma Call (Yes / No) |
| Operative procedure | Anaesthetic Technique (AGP vs non-AGP) | Seniority of Surgeon (Consultants vs juniors) | Comorbidities |
| Six-week mortality (Yes / No) | Post-op complications | Surgery time since admission (hours) | COVID status (Negative / Positive swab) |

### Statistics

All the data were recorded, anonymised and verified by four members of the study group for their accuracy. The data were processed using Microsoft Excel (Microsoft, Washington, USA). Shapiro-Wilk test indicated a normal distribution for patient demographics. Hence, the mean (± standard deviation; 95% CI) were calculated for both age, ASA grade and days to discharge from hospital. Both prevalence or risk and odds ratios were calculated as well as a Fisher’s exact test for statistical significance for categorical data, defined as p≤0.05. Percentages and confidence intervals were rounded off to one decimal place.

### Ethics and permissions

All data points were utilised for routine auditing purposes to reflect departmental activity and service provision without altering clinical care pathways. Each centre contributing data to this study registered their interests with local authority and the auditing or clinical governance departments. No informed consent was required as there was no identifiable data. All data were anonymised at the time of collection and submission. Each patient was assigned a unique identification number which was cross-referenced with the patients’ individual hospital identification or medical record numbers. This cross-referenced list remained internally within the hospital trust computer server handled by the contributing team from each trust. The data was transferred and stored using the NHS.net email server which has been approved for transfer of patient data. Data protection compliance was abided by at all times. The lead center was Imperial College Healthcare NHS Trust where this study was first approved as a clinical audit prior to expanding onto a national scale.

All centres gave permission for the use of their data. This study was assessed using the UKRI/MRC/NHS Health Research Authority Ethics Decision Tool and was considered an ‘audit/not research’; and therefore it was not subject to further ethical review by the NHS Research Ethics Committee (NHS REC).

### Funding

This study required no internal or external funding.

### The collaborative

The COVid Emergency Related Trauma and orthopaedics (COVERT) Collaborative was founded at Imperial College Healthcare NHS Trust. It is currently a member of the COVID Research Group and it has been endorsed by the Royal College of Surgeons of England and Imperial College Healthcare NHS Trust.

### Transparency statement

The Collaborative affirms that this manuscript is an honest, accurate, and transparent account of the study being reported; that no important aspects of the study have been omitted; and that any discrepancies from the study as originally planned have been explained.

### Data sharing

The dataset has been amalgamated for the purpose of this study and will be made available with publication. Anything that may threaten or compromise patient anonymity means that the dataset will not be released or made available within the public domain.

## Results

A total of 4840 clinical encounters took place between the study periods. A total of 172 spinal operations and presentations were excluded from the final analysis. Tables 2 and 3 have categorised these clinical encounters into acute referrals and operative cases between both years.

**Table 2:**
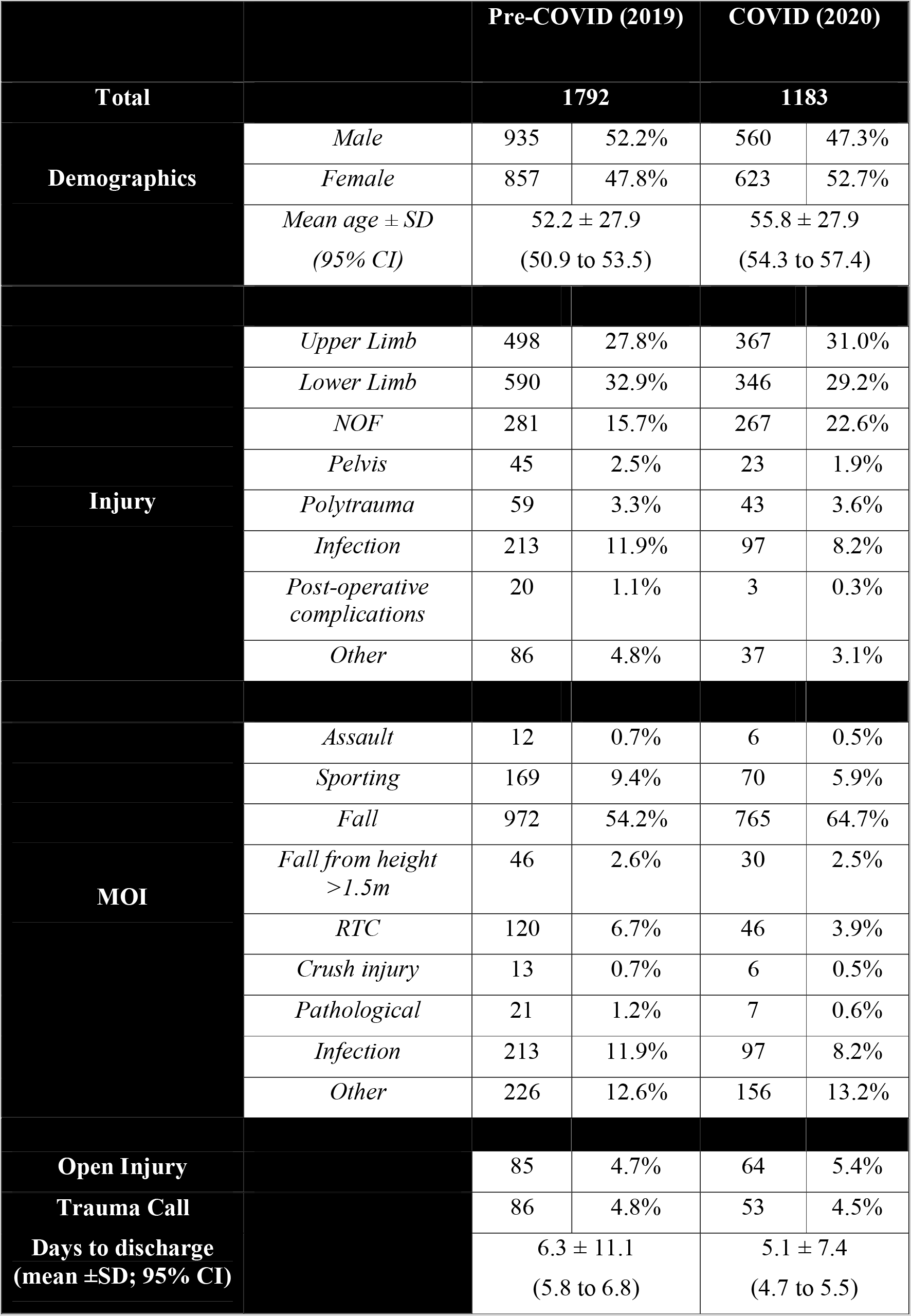
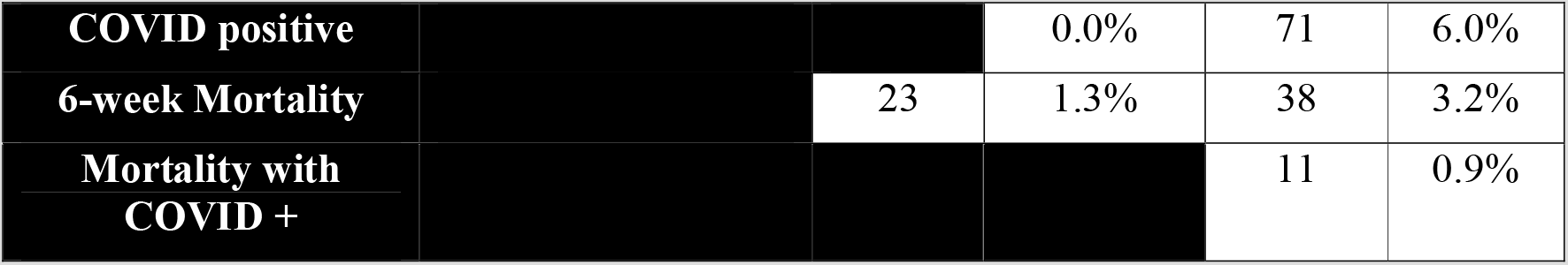
Acute orthopaedic referrals between pre- and post-COVID

**Table 3:**
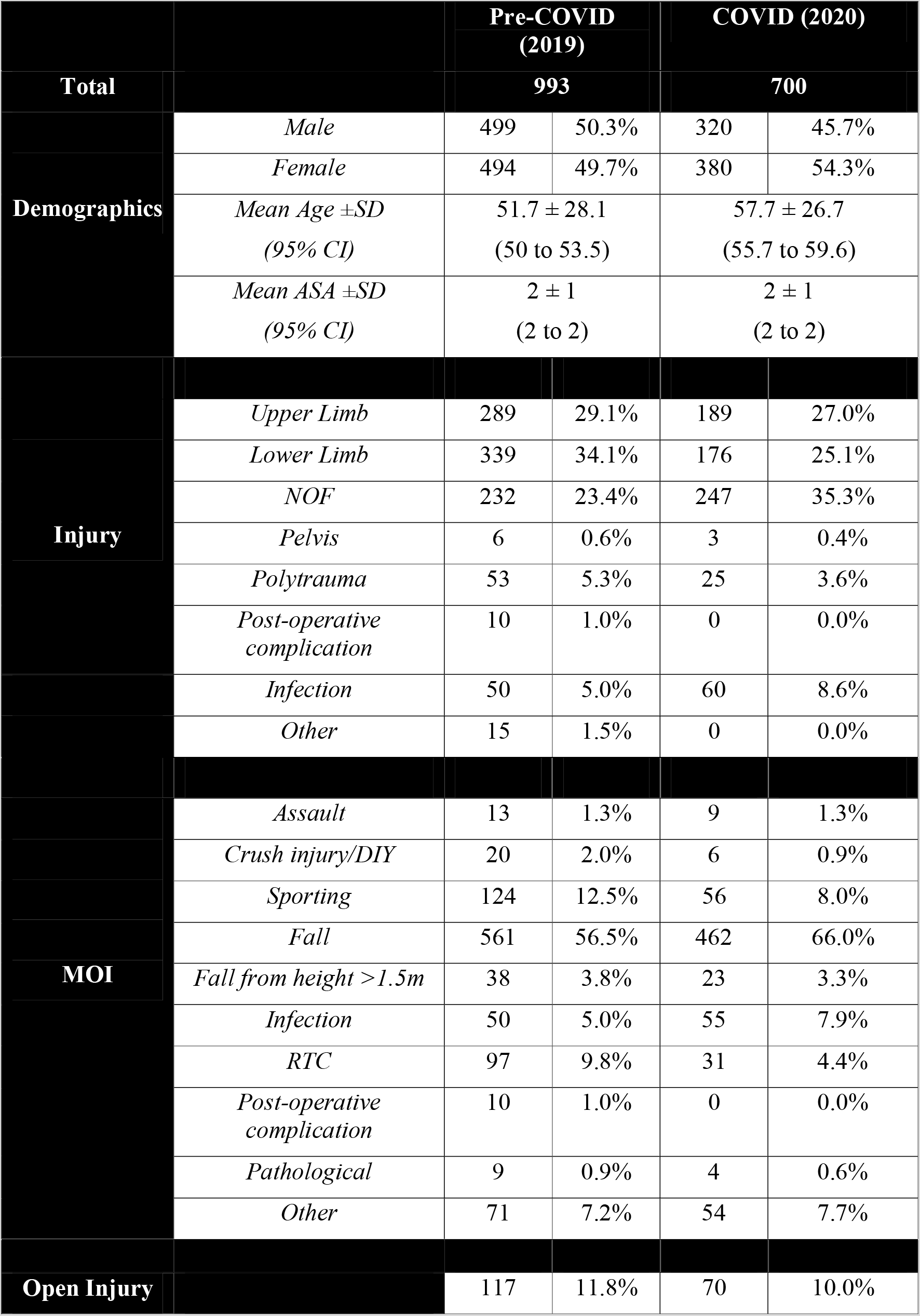
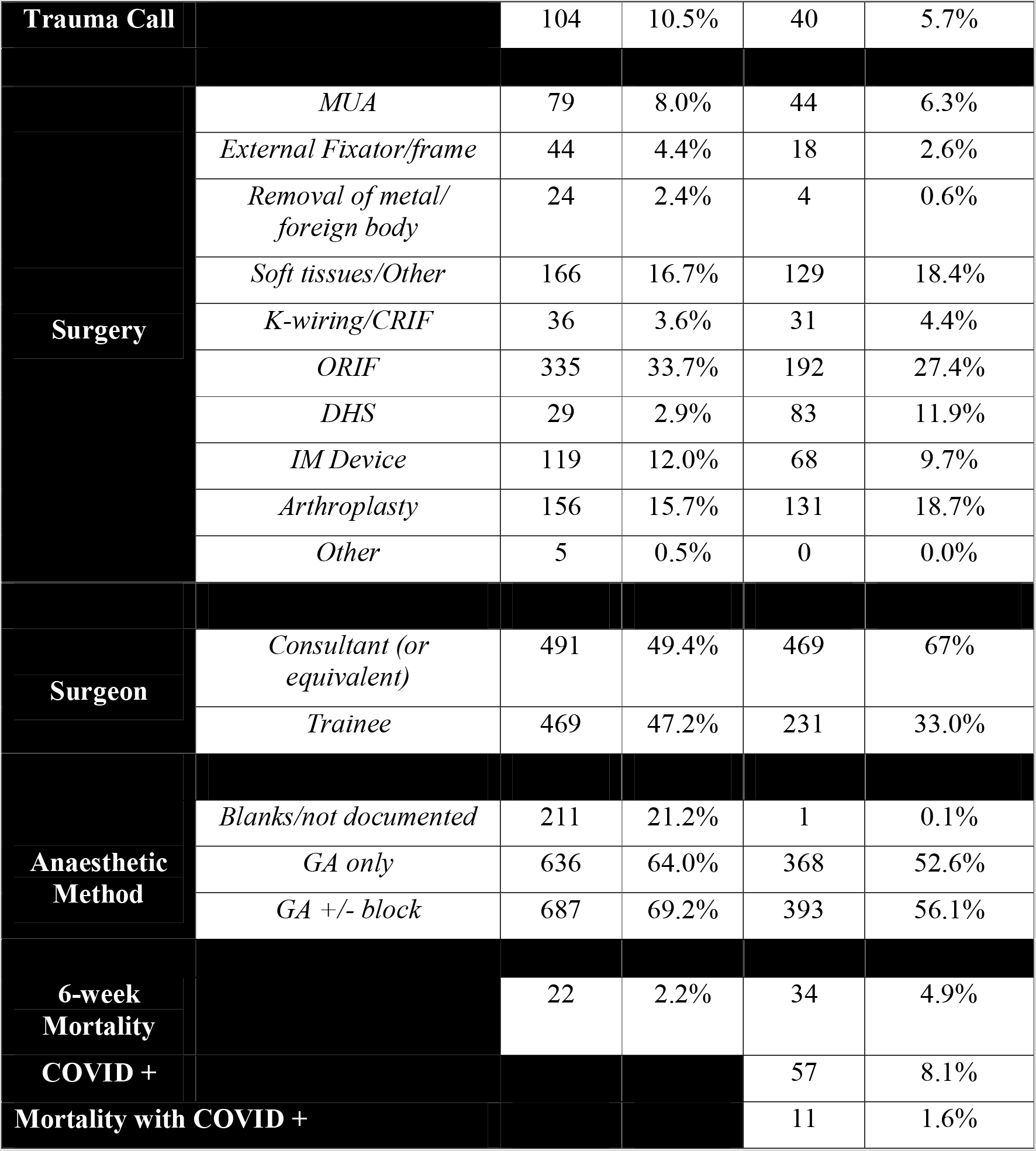
operative caseload between pre- and post-COVID

### Risk (or prevalence) and odds ratios

**Table 4:** Risk (or prevalence) and odds ratios for acute referrals and operative caseloads

|  | Acute referrals |  |  | Operative caseload |  |  |
| --- | --- | --- | --- | --- | --- | --- |
|  | RR<br>(or PR) | OR | p-value | RR<br>(or PR) | OR | p-value |
| Mortality | 2.50 | 2.55 | 0.0005 | 2.19 | 2.25 | 0.004 |
| Mortality due to COVID-related complications vs non-COVID causes | 14.2 | 19.7 | 0.004 | 15.1 | 22.0 | 0.004 |
| General anaesthetic only |  |  |  | 1.22 | 1.61 | 0.00001 |
| General anaesthetic ± block |  |  |  | 1.23 | 1.75 | 0.00001 |
| Consultant-led operation |  |  |  | 1.36 | 2.08 | 0.00001 |
| Open reduction + internal fixation |  |  |  | 0.81 | 0.74 | 0.007 |
| Dynamic hip screw |  |  |  | 2.02 | 2.11 | 0.00001 |
| Removal of metal/foreign body |  |  |  | 0.24 | 0.23 | 0.003 |
| Trauma call |  |  |  | 0.55 | 0.52 | 0.0005 |
| Road traffic accident | 0.58 | 0.56 | 0.001 | 0.45 | 0.43 | 0.00001 |
| Fall (<1.5m) | 1.19 | 1.54 | 0.00001 | 1.17 | 1.49 | 0.0001 |
| Sporting injury | 0.63 | 0.60 | 0.0005 | 0.64 | 0.61 | 0.003 |
| Infection (MOI) | 0.69 | 0.66 | 0.001 | 1.70 | 1.77 | 0.005 |
| Neck of femur (NOF) fracture | 1.44 | 1.57 | 0.00001 | 1.51 | 1.79 | 0.00001 |
| Lower limb (excl. NOF) | 0.89 | 0.84 | 0.04 | 0.74 | 0.65 | 0.0001 |
| Male | 0.91 | 0.82 | 0.01 |  |  |  |
| Other (injury) | 0.65 | 0.64 | 0.0005 |  |  |  |
| Other (MOI) | 1.25 | 1.30 | 0.01 |  |  |  |

Table 4 outlined the risk (or prevalence) and prevalence odds ratios alongside their 95% confidence intervals and statistical significance. The risk ratio is synonymous with the prevalence ratio. Only those factors that were statistically significant within the acute referrals and operative caseloads were included.

Although the expectation was to minimise the use of aerosolising-generating anaesthetic procedures, there was in fact an increased prevalence of using general anaesthesia ± block up to an odds of 75%, perhaps to create a ‘closed circuit’ for the airways. As the anaesthetic methods was not well documented in the pre-COVID era in a fifth of cases, this could have skewed the data as it may have been difficult to extract that data from last year. The odds of a consultant-led operation doubled during the COVID period as a consequence of all elective operations being suspended and more consultants being relocated to trauma theatre and increased pressure within the theatre environments leading to consultant delivered, rather than consultant led care. With respect to surgical procedures, there was a significant reduction in prevalence ratio of open reduction and internal fixation by a fifth and removal of metalwork and foreign bodies by three-quarters, while there was a doubling in dynamic hip screw fixation with an increase by 9% in the COVID era.

With respect to the acute referrals, patients had half the odds of presenting as a trauma call. This could be due to the odds ratios of road traffic accidents, sporting injuries, infection, and lower limb injuries were significantly less (by a third to a half) during the COVID period. Yet, the odds of presenting with a neck of femur fracture and having falls less than 1.5m height increased by more than 50%.

## Mortality

Table 4 indicated that the mortality rate more than doubled significantly for both prevalence (or risk) and odds ratios during the COVID period. This certainly has both statistical as well as clinical significance. COVID-related complications was still responsible for increasing the odds of mortality by 20 to 22 times within all mortalities from both acute referrals and operative cases (as compared to non-COVID causes for all mortality in the year 2019). Table 5 confirmed that the mean age of mortalities across the board were in the elderly patient population with a high mean ASA grade. Males were consistently in the minority, while neck of femur fracture was the modal diagnosis due to falls and persistently in the majority, followed by lower limb injuries. At least 82% of operations were related to neck of femur fractures in which half of all operations during the COVID period involved anaesthetic AGPs. Whereas the mortalities from pre-COVID operations did not have Consultant-led (as primary surgeon) surgery, that increased to three-fifths of all operations conducted during the COVID period. The mean date of presentation to hospital was one week ahead in year 2020 compared to a year ago but the time from admission to mortality differed only by a mean of less than a day in both categories. COVID positive swabs were confirmed in a quarter of acute referrals and in a third of operative cases.

**Table 5:** Patient demographics, details of injuries, mortality rates, and COVID status

|  | *Acute referrals* |  | *Operative casemix* |  |
| --- | --- | --- | --- | --- |
|  | 2019<br>(n=23) | 2020<br>(n=38) | 2019<br>(n=22) | 2020<br>(n=34) |
| Age (mean±SD;<br>95% CI) | 80.2 ± 16.4<br>(73.2 to 87.2) | 77 ± 23<br>(67 to 88) | 83.9±12.2<br>(78.7 to 89.1) | 84.0±13.5<br>(79.4 to 88.5) |
| Male | 9 (39%) | 16 (42%) | 8 (36%) | 15 (44%) |
| ASA (mean±SD;<br>95% CI) |  |  | 3 ± 0<br>(3 to 3) | 3 ± 1<br>(3 to 3) |
| Injury | 12 NOF (52%)<br>4 poly (17%)<br>2 pelvis (9%)<br>2 UL (9%)<br>2 LL (9%)<br>1 other (4%) | 23 NOF (61%)<br>7 LL (18%)<br>3 UL (8%)<br>3 inf (8%)<br>2 other (5%) | 19 NOF (86%)<br>2 LL (9%)<br>1 UL (5%) | 27 NOF (79%)<br>3 UL (9%)<br>3 LL (9%)<br>1 inf (3%) |
| MOI | 20 fall (87%)<br>2 RTA (9%)<br>1 other (4%) | 32 falls (84%)<br>2 other (5%)<br>2 infection (5%)<br>1 RTA (3%)<br>1 sporting (3%) | 21 fall (95%)<br>1 path (5%) | 30 falls (88%)<br>2 inf (6%)<br>1 path (3%)<br>1 other (3%) |
| Open injury | 3 (13%) | 1 (3%) | 1 (5%) | 1 (3%) |
| Trauma call | 2 (9%) | 0 | 0 | 0 |
| Procedure |  |  | 10 hemi (45%)<br>10 IMN (45%)<br>1 DHS (5%)<br>1 MUA (5%) | 13 hemi (38%)<br>10 DHS (29%)<br>5 IMN (15%)<br>3 ST (9%)<br>1 Ex-Fix (3%) |
| AGP anaesthetic |  |  | 9 U/K (41%)<br>7 AGP (32%) | 17 AGP (50%)<br>12 U/K (35%) |
| Consultant-led operation |  |  | 0 | 20 (59%) |
| Date of injury<br>(mean days±SD; | 6/4 ± 11 | 31/3 ± 12 | 6/4 (±12) | 30/3 (±14.2) |
| 95%CI) | (1/4 to 10/4) | (26/3 to 5/4) | (1/4 to 11/4) | (25/3 to 4/4) |
| Time from admission to mortality (mean days±SD; 95%CI) | 10.3 ± 7.5<br>(7.1 to 13.5) | 11 ± 10<br>(7 to 15) | 14.3 ± 10.4<br>(9.8 to 18.7) | 13.8 ± 10.4<br>(10.2 to 17.3) |
| Post-op complication |  |  | 7 (33%) | 11 (32%) |
| COVID status |  | 15 NT/A (39%)<br>8 neg (21%)<br>11 pos (29%)<br>4 U/K (11%) |  | 11 pos (32%)<br>9 NT/A (26%)<br>8 U/K (24%)<br>6 neg (18%) |

**Key:** *LL: lower limb, UL: upper limb, path: pathological, AGP: aerosolizing generating procedure (anaesthetic), U/K: unknown, hemi: hemiarthroplasty, NOF: neck of femur fracture, MUA: manipulation under anaesthesia, poly: polytrauma, inf: infection, IMN: intramedullary nailing, ST: soft tissue; RTA: road traffic accident, NT/A: not tested/asymptomatic, neg: negative, pos: positive*

Taking into account that COVID was a peri-operative complication since patients may have been symptomatic with COVID manifestations pre-operatively but only had the swab results return with a positive finding either pre- or post-operatively; the commonest post-operative complication in the COVID period was a hospital-acquired pneumonia but with negative COVID swab results or the decision not to test at all. The second most common postoperative complication in the year 2020 was extra-pulmonary sepsis. This was seen in table 6. The proportion of post-operative complications were similar in both years.

**Table 6:** Post-operative complications for both years

| 2019 |  | 2020 |  |
| --- | --- | --- | --- |
| 1 | Acute deterioration | 11 | COVID positive<br>(pre or post-op) |
| 1 | Bowel obstruction | 5 | HAP<br>(COVID swab negative or not tested) |
| 1 | Cellulitis | 4 | Extra-pulmonary sepsis |
| 1 | NSTEMI | 3 | Delirium |
| 1 | HAP | 2 | AKI |
| 1 | Anaemia | 2 | Anaemia |
| 1 | Cardiac arrest | 1 | Upper GI bleed |
|  |  | 1 | Cardiac arrest |

**Key:** NSTEMI: non-ST elevated myocardial infarction, HAP: hospital-acquired pneumonia, AKI: acute kidney injury, GI: gastrointestinal

Table 7 focussed on the total number and nature of comorbidities within the mortality groups. Multiple contingency chi-square test was insignificant for both number of comorbidities and individual comorbidities between both years, except for cardiovascular and cerebrovascular disease in acute referrals. This was corroborated by the COVIDSurg publication^12^ which confirmed a significant association of mortality with myocardial infarction and congestive heart failure. However, hypertension and stroke/transient ischaemic attacks were not significantly associated. In our study, all cardiovascular diseases (including peripheral vascular, arrhythmias, hypertension, heart failure, myocardial infarction and acute coronary syndromes) were combined with cerebrovascular diseases (consisting of strokes and transient ischaemic attacks). Unlike their study, our study did not find a significant association with chronic kidney disease, chronic obstructive disease (which included asthma) and dementia in all mortalities during the 2020 timeframe regardless of the COVID status. The differences may stem from that their study looked at the comparison of mortality rates within the same cohort during the COVID era, whereas this study is sub-analysing the entire mortality cohort on its own to observe for specific associations and risks.

**Table 7:** Mortality analysis for acute referrals between both years

|  | *Acute referrals* |  | *Operative casemix* |  |
| --- | --- | --- | --- | --- |
|  | **2019**<br>**(n=23)** | **2020**<br>**(n=38)** | **2019**<br>**(n=22)** | **2020**<br>**(n=34)** |
| Diabetes mellitus | 6 (26%) | 7 (18%) | 1 (5%) | 7 (21%) |
| Cardiovascular/cerebrovascular disease | 12 (52%) | 31 (82%)<br>(p=0.01) | 12 (55%) | 27 (79%) |
| Chronic obstructive pulmonary disease/asthma | 6 (26%) | 9 (24%) | 5 (23%) | 7 (21%) |
| Renal disease | 2 (9%) | 10 (26%) | 2 (9%) | 10 (29%) |
| Dementia | 8 (35%) | 16 (42%) | 6 (27%) | 12 (35%) |
| Cancer | 2 (9%) | 6 (16%) | 4 (18%) | 7 (21%) |
| Autoimmune | 0 | 0 | 0 | 2 (6%) |
| None | 2 (9%) | 2 (5%) | 0 | 1 (3%) |
| ***Number of comorbidities*** |  |  |  |  |
| 0 | 2 (9%) | 2 (5%) | 0 | 1 (3%) |
| 1 | 9 (39%) | 10 (26%) | 5 (23%) | 12 (35%) |
| 2 | 9 (39%) | 15 (39%) | 10 (45%) | 9 (26%) |
| 3 | 1 (4%) | 10 (26%) | 3 (14%) | 7 (21%) |
| 4 | 2 (9%) | 2 (5%) | 1 (5%) | 4 (12%) |
| 5 | 0 | 0 | 0 | 1 (3%) |
| Unknown / not documented | 1 (4%) | 0 | 2 (9%) | 0 |

## Survival probability

A six-week Kaplan-Meier survival probability analysis for mortalities between both years was plotted in figure 1.

**Figure 1:**
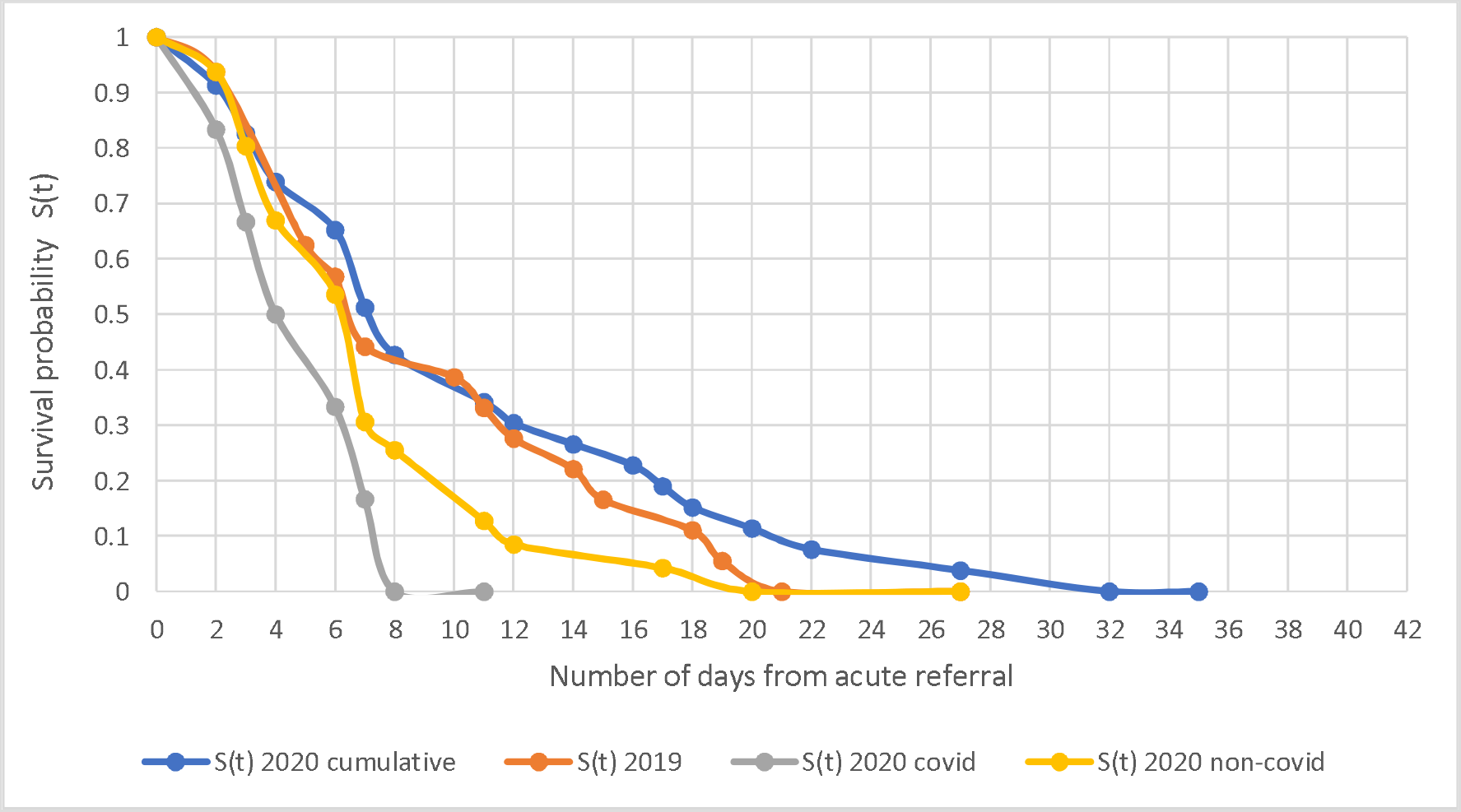
Six-week Kaplan-Meier survival probability analysis for mortalities between preand post-COVID for acutely referred from the Emergency Department

There were similar patterns of survival probability between both cohorts (i.e. 2019 vs 2020 cumulative). However, the lowest survival probability and the shortest timeframe were observed in the confirmed COVID positive cohort as seen in figure 1. This may be due to the most vulnerable patient profile. 8 (72.7%) patients had femoral trauma, most being neck of femur fractures, distal femur fracture and a dislocated hip hemiarthroplasty post-fracture. Other patients presented with septic arthritis, post-operative complication and knee swelling; yet every patient also suffered from multiple comorbidities including those leading to immunosuppression as seen in tables 5 and 7. Although these patients were prioritised in the Emergency Department and recognised for their poor physiological reserve, due to the stresses of the acute and emergency services, these patients may have had to wait longer to be treated acutely and appropriately admitted.

**Figure 2:**
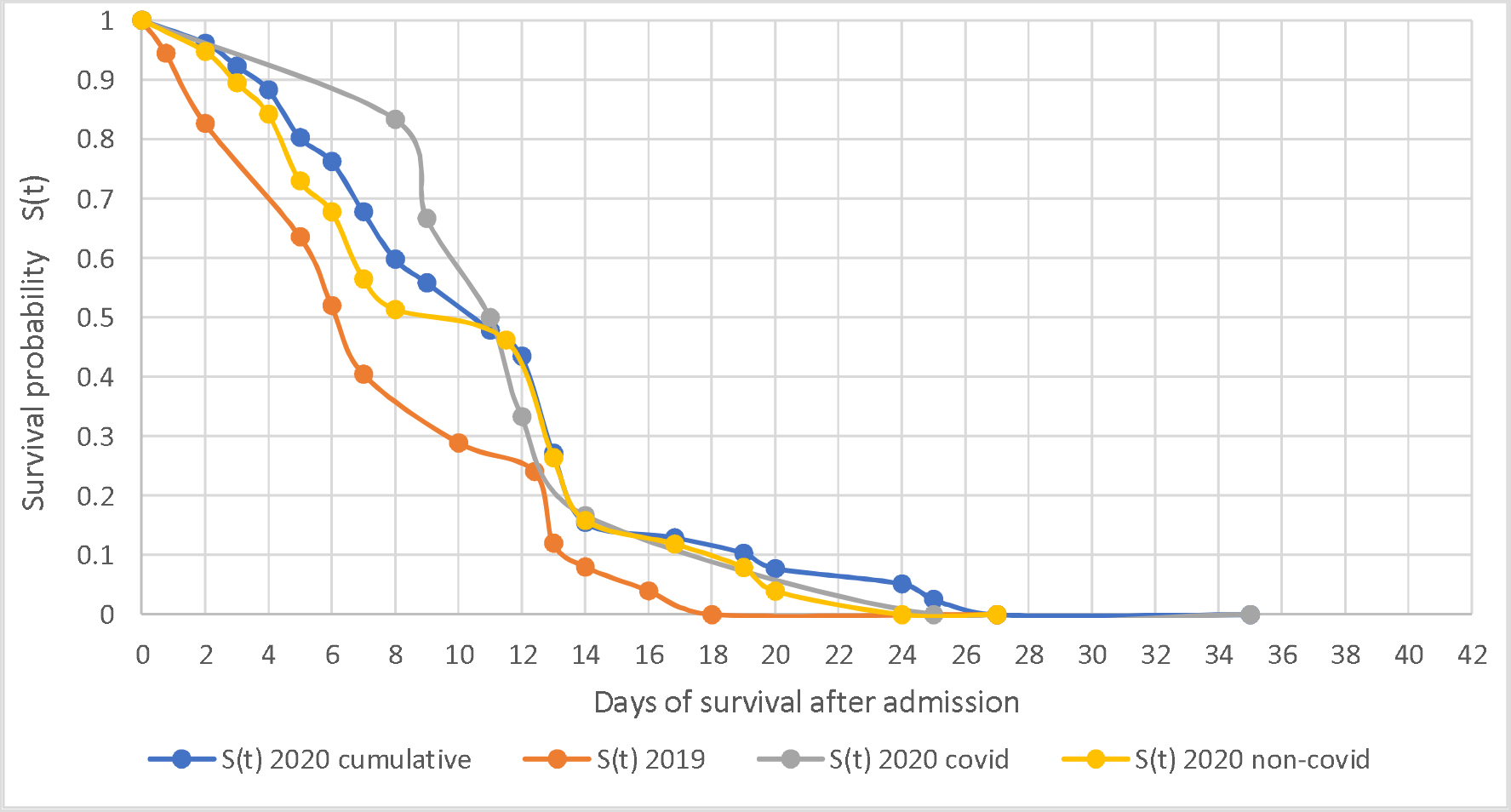
Six-week Kaplan-Meier survival probability analysis for mortalities between preand post-COVID for those undergoing surgery

Unexpectedly, there was a reversal of trends observed for the six-week Kaplan-Meier survival analysis once admitted and operated on in figure 2. Mortalities within the pre-COVID period had the lowest survival probability compared to the post-COVID cohort. The COVID positive mortalities were observed to have the highest survival probability 11 days prior to converging with those mortalities without COVID symptoms. This was most likely to be due to multifactorial factors. During the pandemic, wards were ring-fenced to host confirmed COVID positive patients with a heightened care of nursing, medical cover and personal protective equipment. Prior to the onset of a possible vaccination to counteract the virus, symptomatic management and shielding were the mainstay treatments for COVID positive patients. None of these patients were stepped up to the intensive treatment unit due to being categorised as high risk stratification for mortality based on age and extent of comorbidities. If these ‘at risk’ patients were symptomatic with the virus, then aggressive pre-operative optimisation would occur. Since 91% (n = 10) of COVID positive patients had sustained a neck of femur fracture, the National Hip Fracture Database best practice tariff of operating within an ideal 36-hour window set by the Royal College of Physicians was suspended until the patient was stabilised. All hip fracture patients in this cohort were operated on and had dedicated orthogeriatric input commencing from hospital admission. Hence the early peri-operative period and surgery encompassed within the 10-day period post-admission. Moreover, neck of femur fractures are recognised as a pre-terminal illness and are known to carry a high risk of mortality in the first month which is trebled in the first year after the injury.^13^

## Discussion

### Statement of principal findings

There was a significant difference between pre- and post-COVID periods at its ‘golden peak weeks’. The alternative hypothesis was not rejected with respect to prevalence of (i) acute orthopaedic trauma referrals (reduced by 34%), (ii) surgical interventions (reduced by 29.5%), (iii) anaesthetic aerosolising generating procedures, (iv) mortality rates (more than doubled in the COVID period), and (v) survival probability between pre- and post-COVID eras. The 34% reduction in acute trauma referrals is in keeping with previous single centre studies performed in the UK with results ranging between 33-50%.^10,11^ As described in these previous studies we would attribute the overall reduction of trauma workload to be due to reduction in travel and outdoor activities during the national lockdown. There was a significant decline in the odds of trauma calls, road traffic accidents, sporting injuries and lower limb fractures. Conversely, there was a significant rise in the odds of neck of femur fractures, falls, the use of anaesthetic AGP and consultant-led operations. Since the aetiology of neck of femur fracture are often low energy falls in the home environment, it is not unexpected to observe a consistency of neck of femur fractures in the elderly and the vulnerable during lockdown. Odds of falls may have increased due to prodromal symptoms and clinical manifestations of COVID.

### Mortality rates

Mortality rates significantly doubled for both prevalence (or risk) and odds ratios during the COVID-19 period and a third of those deaths had a positive COVID-19 diagnosis. Those with neck of femur fractures remain at greatest risk of mortality and there have been further studies evaluating the risk of COVID-19 on this inherently high risk cohort.^14,15^ Comparatively, the COVIDSurg Collaborative observed a 30-day mortality rate of 28.8% (p< 0.0001) of Orthopaedic patients who underwent surgery (both elective and trauma) within the first quarter of the year.^12^

As lockdown measures in the UK and globally eases and the incidence of trauma returns to pre-lockdown trends, it is imperative that we understand the true increased risk of mortality in the acute trauma patient during the COVID-19 era. A recent publication by Kader et al.^16^ has suggested that the rate of mortality from COVID-19 for elective Orthopaedic patients is low; yet this is the first nationwide study to quantify mortality risk for trauma patients. Trauma procedures due to the nature of the injuries are necessary and time-critical, and nobody can afford to postpone trauma care even during a global pandemic.^17^ With an overall mortality risk in 2020 doubled that of 2019, clinicians need to counsel patients presenting with acute orthopaedic trauma of the increased risk in the COVID-19 era, especially for those identified as increased risk stratification with multiple underlying comorbidities, elderly and frailty. With the ongoing risk of a second wave and resurgence of COVID-19 cases on top of the inevitable winter pressures, this data is of critical importance in the risk management, decision-making and policymaking of trauma patients both in the UK and across the globe.

### Strengths and weaknesses of the study and in relation to other studies

This was the first representative observational study of the UK looking into the impact of COVID-19 pandemic on general Trauma and Orthopaedic surgical speciality. Studies thus far have only shed light on local scales or cross-speciality.^10–12^ Weaknesses included loss of data points which have been accounted for in the tables (i.e. tables 2, 3, 5 and 7 labelled as unknown). However this did not affect the final analysis of data points.

### Limitations and future research

Albeit the largest study on a national scale within this speciality, it is vital to continue exploring the impact of the pandemic on a larger scale. Ideally, more secondary care providers consisting of district general hospitals and major trauma centres ought to be recruited to submit data. Data ought to be submitted during the peak of the pandemic as well as at various time intervals as the lockdown measures continue to ease and there is more freedom of movement for the public but the ever-present risk of a second wave.^18^

## Conclusion

This was the first national representation of the impact of COVID-19 pandemic on acute Orthopaedic trauma referrals and mortality between mid-March to end-April, representing the ‘golden peak weeks’ during the lockdown. The mortality rate for acute referrals, as well as those undergoing operative intervention, more than doubled in odds when compared to the same time interval one year ago. The majority of mortalities consisted of the elderly with neck of femur fractures and cardiovascular and/or cerebrovascular diseases. This study will aid clinicians in counselling trauma patients of the increased risk of mortality during the era of COVID-19 and also aid in both healthcare infrastructure, resource allocation, decision-making and policymaking as we continue to battle with the pandemic.

